# Type 1 Fuzzy Logic Classification of Pain Severity (Pain Assessment)

**DOI:** 10.1101/2023.01.23.23284915

**Authors:** Linnaeus T. Bundalian, Rhendell John M. Pariño, Rionel B. Caldo

## Abstract

Fuzzy logic has given man a vast range of opportunities in different field of practice. Fuzzy logic applications are often used in automated systems, robotics and even in medicine. Medical applications of fuzzy logic has greatly improved some gray areas in medical practices especially when considering factors that has no direct unit of measurement to define its degree or magnitude and this includes pain assessment. Assessing pain has become a challenge for medical practitioners since the degree of its severity varies among people. There are number of tools to assess the severity of pain, but none of these have described a constant scale for each corresponding degree of severity. This paper applies the concepts of fuzzy logic in assessing the severity of pain. The goal of the study is to come up with a type 1 fuzzy logic classification of pain assessment in Excel VB macro program platform.

## 1. INTRODUCTION

There are number of difficulties encountered in decision-making and assessment for medical and healthcare practice. The challenge extends mostly among decisions and diagnosis that involve the patient’s self-report intensity [1]. These intensities are dependent on human intuition and reasoning, hence it may vary for every individual and defining such assessment involves considerable uncertainties. One of the most difficult factors to assess is pain. Pain is an unpleasant feeling often caused by intense or damaging stimuli [2], which can be assessed and determined using sensorial information [1]. Different pain assessment tools have been developed to address the need for a definite diagnosis of its severity but still pain is a perception-based stimuli and these tools might not be precise in giving results. The proponents have decided to assess the parameters being observed on different pain assessment tools to come up with generalized results in the context of tools under consideration. In this study, the proponents will develop a fuzzy-based assessment system, which can assess the pain severity by referring to the parameters derived from different pain assessment tool, namely Wong Baker Facial Grimace Scale, Activity Tolerance Scale and FLACC.

Pain is considered as the fifth vital sign for representing basic bodily functions and health quality of life [3][4]. Pain is a fundamental element in healthcare [5]. Unidimensional or multidimensional characteristics are often employed in measuring pain depending on the objectives and interests achieved in the course of diagnosis but still there is no standard approach that allows the objective, external observation other than the patient’s self-report. Self-report is a preferable pain indicator and necessary for correct evaluation of pain manifestation but still there is a need for a common language to translate pain between the patient and medical or healthcare professionals; these are the pain measurement scales or pain assessment tools [6]. In this study, the proponents will consider three of pain assessment tools: Wong Baker Facial Grimace Scale, Activity Tolerance Scale and FLACC. These subjectivity of pain will be determined by using fuzzy sets as derived from the tools under consideration. The proposed approach secures the achievement of the determination of the presence, intensity and significance of pain.

***Type 1 Fuzzy Logic Classification for Pain Assessment*** is an attempt to develop a fuzzy-based system, which can collect, analyze and assess different parameters being observed in pain assessment tools automatically. The system to be developed is an Excel VB Macro program. The results of the program is to be compared to the results of a Fuzzy-based program made in MatLab.

### 1.1. Objectives

The prime objective of this study is to apply fuzzy logic in assessing pain severity.

Specifically, this study aims to:

1. describe the fuzziness of pain severity
2. develop a fuzzy-based pain assessment system
3. monitor the severity of pain for all the users
4. test and evaluate the performance of the developed fuzzy system

## 2. FUZZY LOGIC SYSTEM

### 2.1. Fuzzy Description of Pain Severity

Fuzzy Rule-based system is used to assess the severity of pain. In a fuzzy rule based system, the proponents enumerated a list of pain severity parameters to represent pain severity classification in the form of rules. The rules defined by the system has its attribute name which are FLACC, Facial Grimace and Activity Tolerance and its corresponding linguistic values like No Pain, Mild Pain, Moderate Pain, Severe Pain and Extreme Pain. In the course of study, since the proponents have considered activity tolerance and facial grimace as a critical parameter, the said parameters were excluded from FLACC (Face-Legs-Activity Tolerance-Cry-Consolability) thus considering legs, cry and consolability only in FLACC parameters.

#### A. Face Pain Scale

The faces in the scale portraits how severe the pain is using visual facial expression which ranges from no pain to worst pain possible. The Wong Baker’s Facial Grimace uses six scales in total; the proponents have modified the scale in reference to the VASOF (amalgamation of Visual Analog Scale and Scale of Faces) [7].

#### B. Activity Tolerance Scale

The Activity Tolerance scale describes the pain level in terms of what the patient is capable of. The scale is also modified to come up a fix range for the scales that will fit to the other parameter ranges.

#### C. FLACC

FLACC assessed pain by considering the parameters which are face, legs, activity, cry and consolability. The proponents have considered legs, cry and consolability as FLACC parameters since face and activity are critical elements for the study.

### 2.2. Fuzzy Method

The proponents have identified the pain assessment tool and scale as a reference for the input parameters of the study. The proponents had collected relevant data for references to provide and generate data to be subjected under analysis. Through the data which have been gathered, the proponents have derived the following input parameters: Legs, Cry and Consolability for FLACC parameter, while FLACC parameter will then be used as an input parameter together with Facial Grimace and Activity Tolerance for the determination of pain severity.

### 2.3. Fuzzy Rule Based System

A set of fuzzy rules was constructed for pain assessment classification which are No Pain, Mild Pain, Moderate Pain, Severe Pain and Extreme Pain. Each rule with its propositions is ANDed together to come up with some consequences [6].

A hierarchical structure was constructed for the classification of pain severity, *Refer to figure 2.1*. The following are the sample rules stored at the different hierarchical levels of the structure:

**Figure 2.1.**
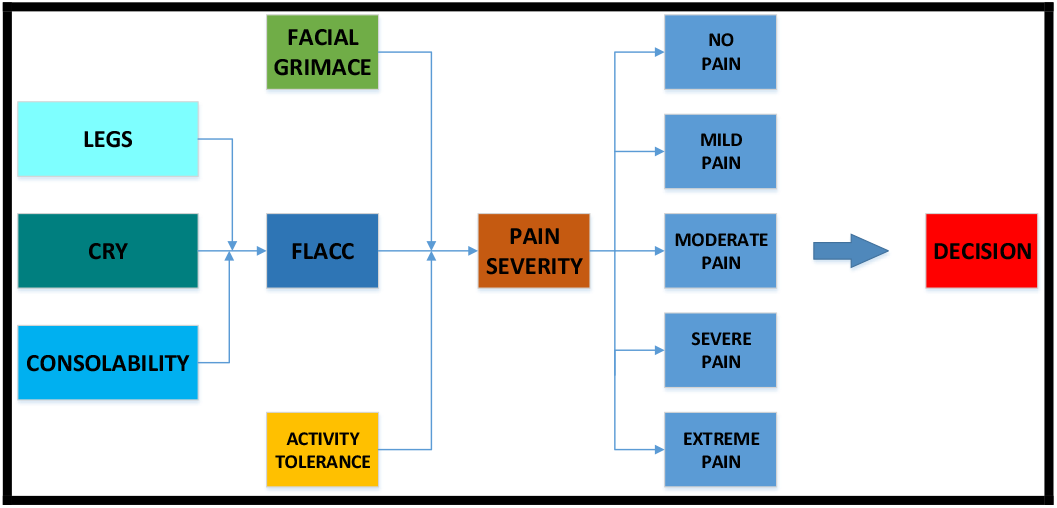
Hierarchical Structure for Pain Severity Classification

*If LEGS is <Normal> and CRY is <normal> and CONSOLABILITY is <Normal>*

*Then FLACC is <No Pain>*

The rule at the last level could be *If FLACC is <No Pain> and Facial Grimace is <No Pain> and Activity Tolerance is <No Pain>* ***Then Pain Severity is <No Pain>***

In this research, the precedence graph has been built analyzing the sequence or flow of data involved in assessing pain severity. In a precedence graph each task is represented by a node, node arrows connect a task to its corresponding subtasks.

### 2.4. How it Works: Type 1 Fuzzy Logic Classification of Pain Severity

The proponents illustrate the use of fuzzy logic in medical and healthcare assessment with fuzzy linguistics for pain severity. They make use of fuzzy classification to characterize pain severity. The process of defining pain severity involves a great deal of uncertainty. The user of the program will provide the necessary inputs for the critical parameters which will be normalized to adjust to the system scale and ranges. A built in pie chart is placed on a sheet for monitoring purposes. The number of patients or users that experience a particular pain scale rating is being recorded and placed on a chart.

## 3. METHODOLOGY

### 3.1. Design of Fuzzy Logic for the pain severity assessment

The basic process of designing a fuzzy logic for the pain severity or assessment system involves five (5) steps:

a. **Formulating the problem and selecting the input and output variables state**. For this pain severity/ assessment system the inputs to the fuzzy system are the FLACC, Facial Grimace and Activity Tolerance. The manipulated variable is produced (scored) and sent to the pain severity(output).
b. **Selecting the fuzzy inference rules**. This generally depends on human experience and trial-and-error. The inference rule is selected based the information gathered and derived from existing pain assessment tool. The values are averaged and rounded off to fit linguistic terms.
c. **Designing fuzzy membership functions for each variable**. This involves determining the position, shape as well as overlap between the adjacent membership function, as these are major factors in determining the performance of the fuzzy logic. In defining membership function, geometric shapes such as triangular, trapezoidal, etc are used. The selection is dependent on the expert’s knowledge and understanding of the process (Aprea et al., 2004; Ross, 2004). In this study, the proponent considers triangular membership functions.
d. **Performing fuzzy inference based on the inference method**. Smoothness of the final control surface is determined by the inference and defuzzification methods.
e. **Selecting a defuzzification method to assess the pain severity**. The choice of the defuzzification method determines to a large extent the “quality” of assessment. Hence, it must be chosen carefully. In this case defuzzification is done by calculating the center of gravity and the output is produced through averaging techniques.

In summary, a fuzzy decision is the result of weighing the evidence and its importance in the same manner that humans make decisions. Fuzzy logic reflects humanlike thinking where the human can deduce an imprecise conclusion from a collection of imprecise premises [8].

In this study, the proponents used Sugeno method for the following compelling reasons: a) it is simple, b) it is suited to system whose output is constant or linear in nature.

### 3.2. Input/Output Membership Functions

There are three inputs into the pain assessment fuzzy system, the FLACC, Facial Grimace and Activity Tolerance. There are five membership functions describing each parameters. Each function is labeled No Pain, Mild Pain, Moderate Pain, Severe Pain, and Extreme Pain and is shown in *figure 3.2a*. The FLACC parameter requires three inputs, legs, cry and consolability whose membership functions are No Pain, Mild Pain and Severe Pain. The membership functions are identically labeled for better analysis. The output of assessing the severity of pain are No Pain, Mild Pain, Moderate Pain, Severe Pain, and Extreme Pain. These functions represent a degree of a binary value, 1 being the highest and 0 being the lowest, and are shown in *figure 3.2b*.

**Figure 2.2.**
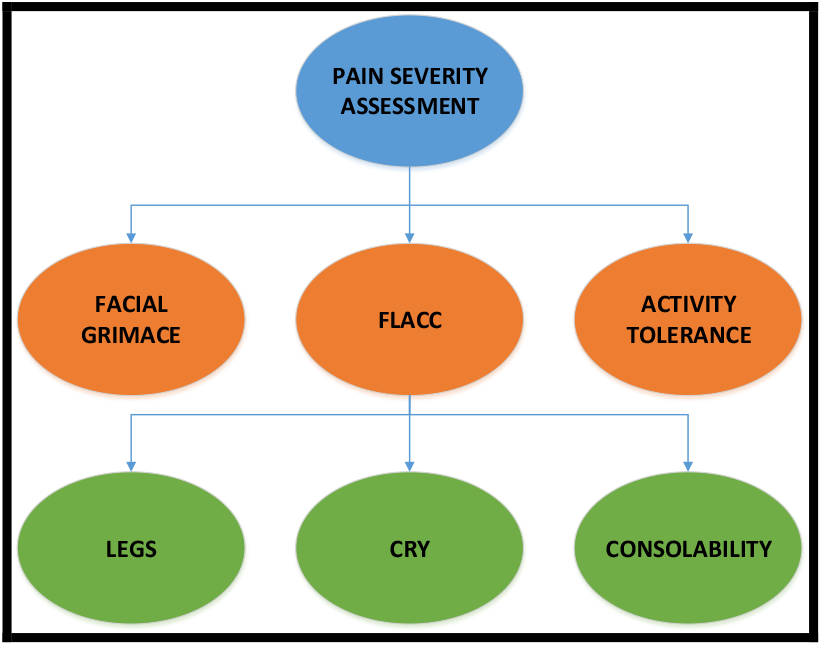
Precedence Graph of Pain Severity Assessment

**Figure 3.1a.**
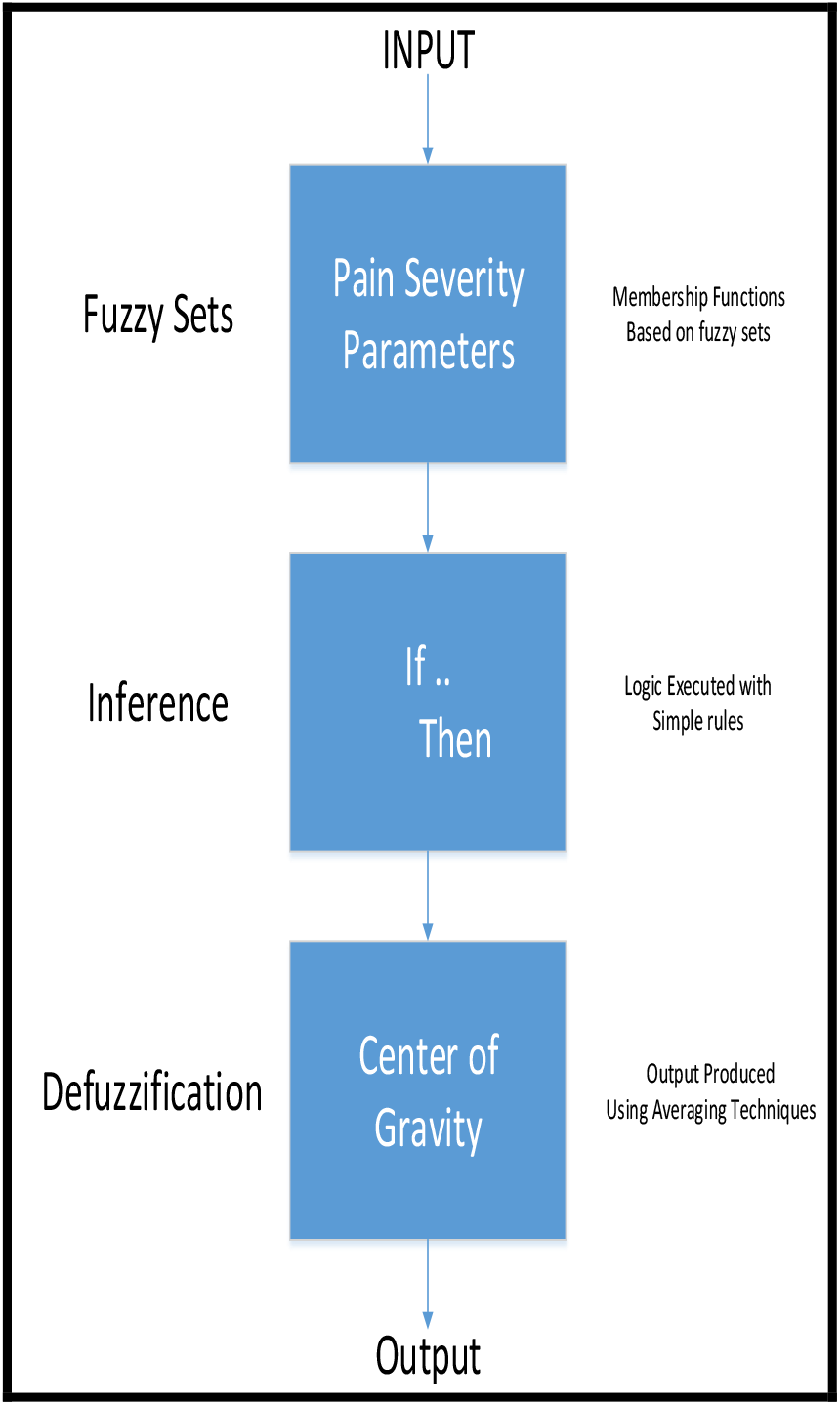
Fuzzy Logic Control Process

**Figure 3.1b.**
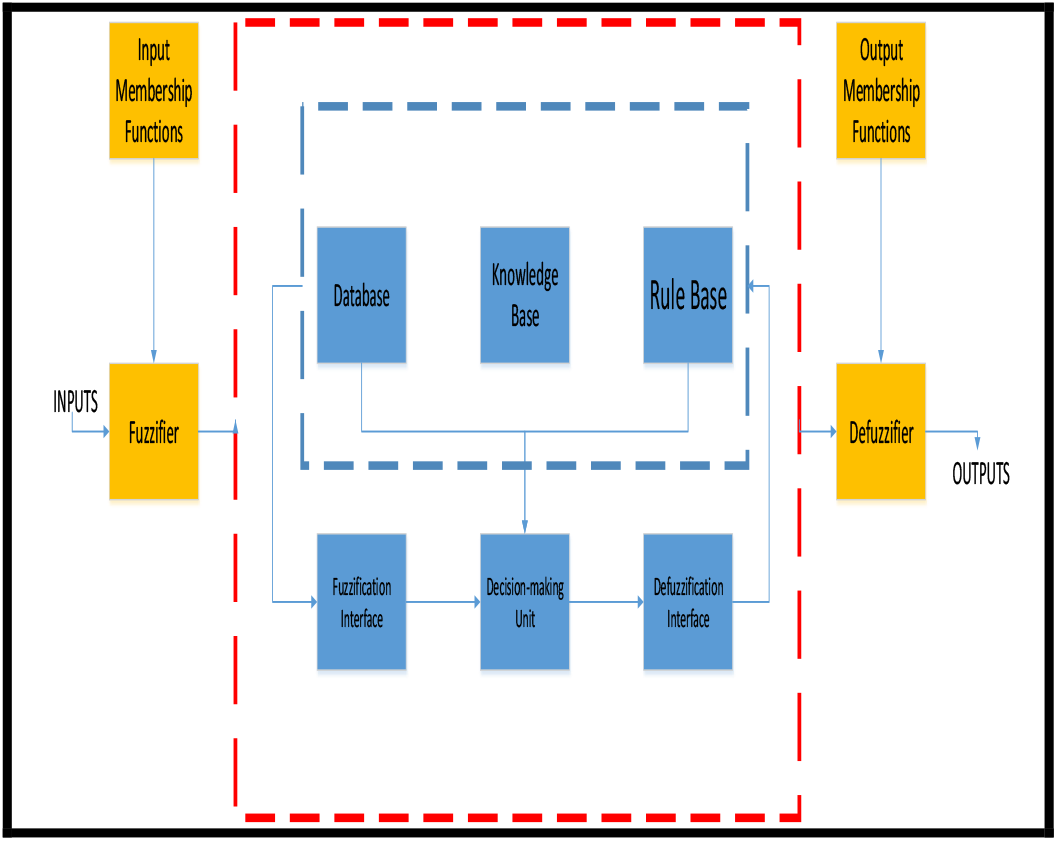
Fuzzy Inference System (FIS) Configuration

**Figure 3.2a.**
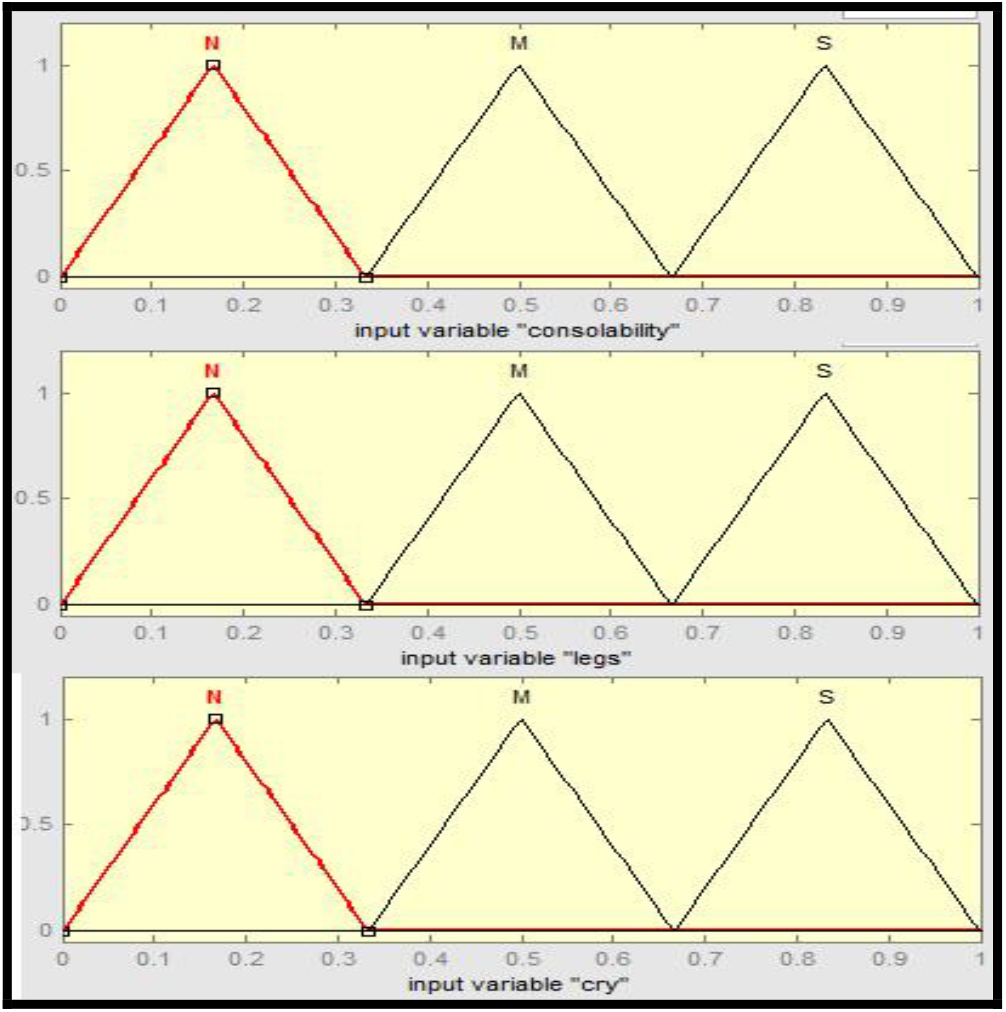
Membership Functions for FLACC: a) Consolability; b) Legs c) Cry

**Figure 3.2b.**
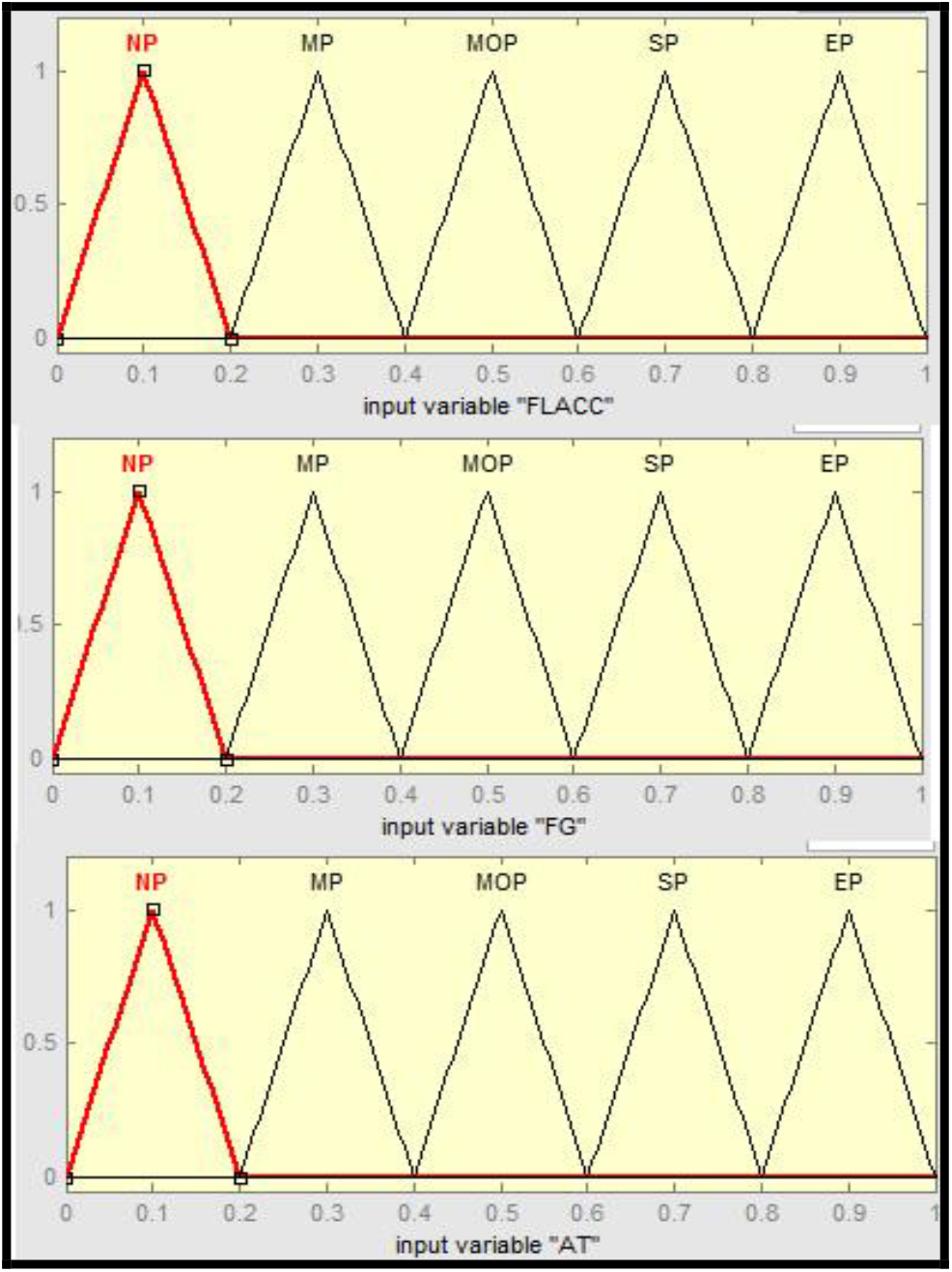
Membership Functions for Pain Severity: a) FLACCs; b) Facial Grimace; c) Activity Tolerance

### 3.3. Project Description

#### a) Properties of the Project

Pain Assessment (Pain Severity) System is a Fuzzy-based centralized macro program. This software consolidates and assesses significant pain severity parameters derived from the assessment tools under consideration. (**FLACC, Facial Grimace and Activity Tolerance**). The key property of this innovation is that, it promotes advanced assessment of pain severity with high degree of certainty and accuracy. This software is flexible in changing data inputs. It is said to be user-friendly since it is easy to use and is built with cutting edge design features and animations. The program is also capable of monitoring the number of patients or users experiencing a particular pain scale rating.

#### b) Functions of the Project

To understand each functions of the project, the proponents tabulate them. Refer to table 3.3 below for the Functions of Pain Assessment System.

**Table 3.3.** Functions of Pain Assessment System

| Item No. | Functions | Definitions/Description |
| --- | --- | --- |
| 1 | Centralized | Storage of the pain severity parameters are stored in a single PC |
| 2 | Fuzzy-Based | Decision-making methodologies are replaced by fuzzy-based ruling system |
| 3 | Faster Data Updates | Real-time and automatic updating of data upon encoding |
| 4 | Data Monitoring | Data are being monitored upon encoding. A pie chart is generated to monitor the number of users or patients experiencing a particular pain scale rating |
| 5 | Fool-Proofing Methodologies | To avoid occurrences of bugs like data type mismatch and insufficient values for inputs, the system has built in pop-ups, disabling/enabling properties and message boxes |
| 6 | User-Friendliness | The system was made under Visual Basic platform and has a design that can be easily understood by its user |

Table 3.3 shows four features of the system. These features are deemed to be helpful in providing reliable and effective data results.

#### c) Tools and Methodologies of the Project

Figure 3.3a is the system main page. It serves as a platform for inputs of data samples. This page displays the summary of the inputs provided by the user.

**Figure 3.3a.**
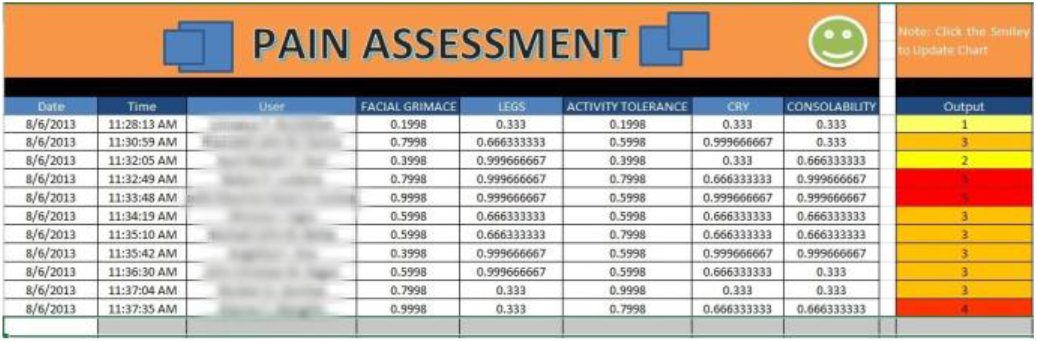
System Main page

**Figure 3.3b.**
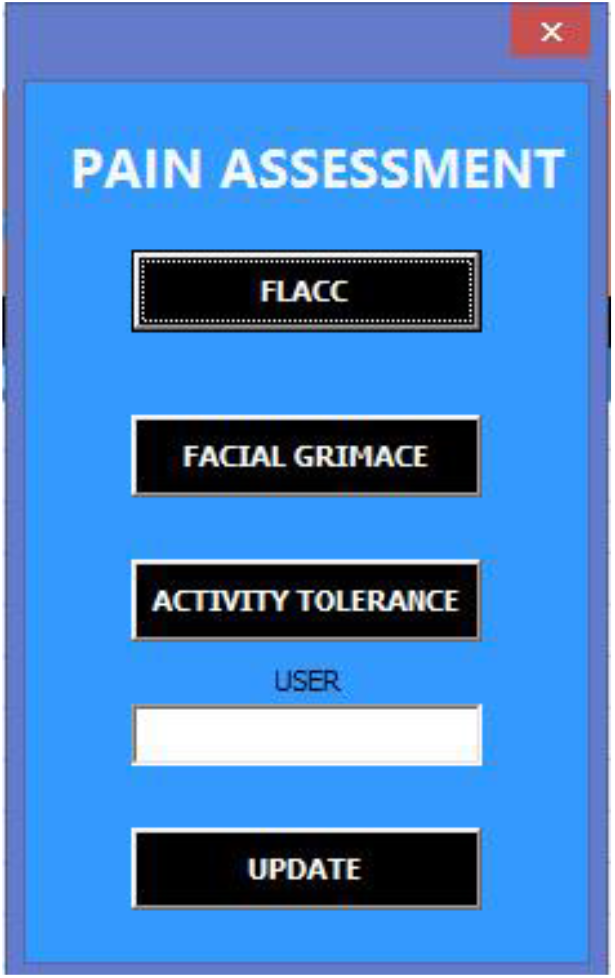
System Homepage

**Figure 3.3c.**
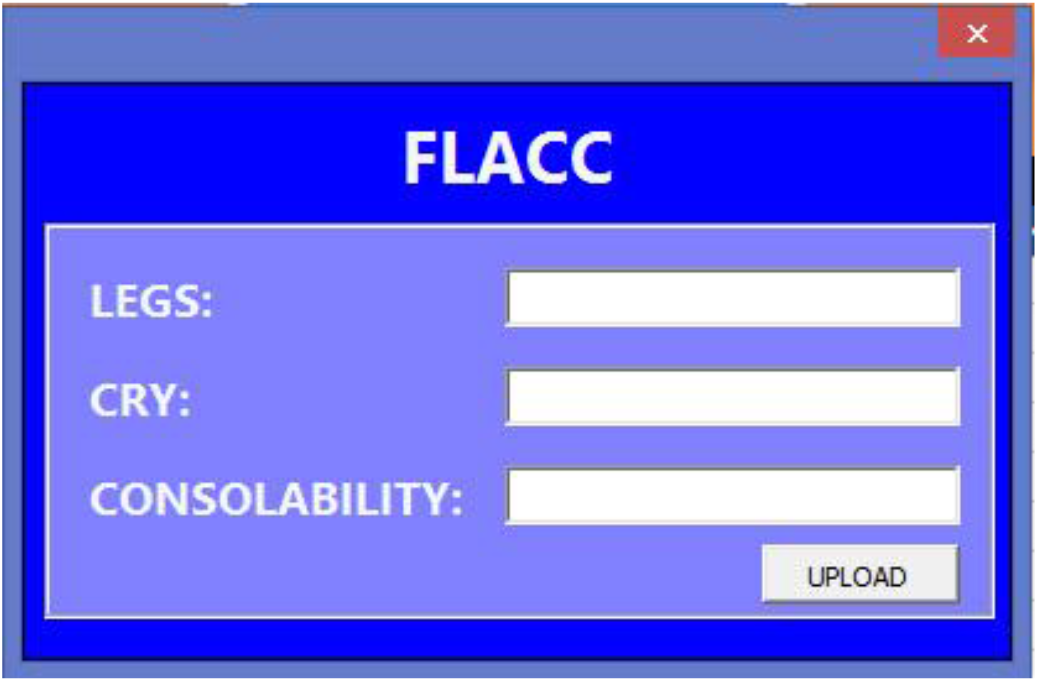
FLACC User Form

**Figure 3.3d.**
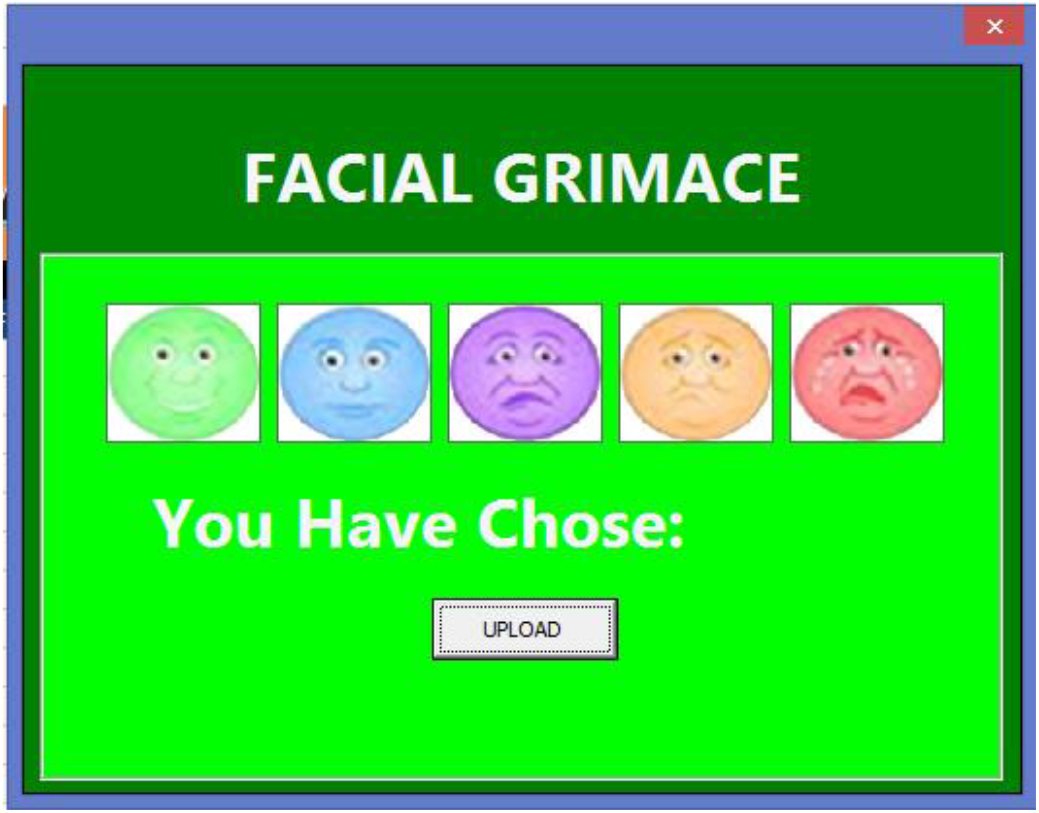
Facial Grimace User Form

**Figure 3.3e.**
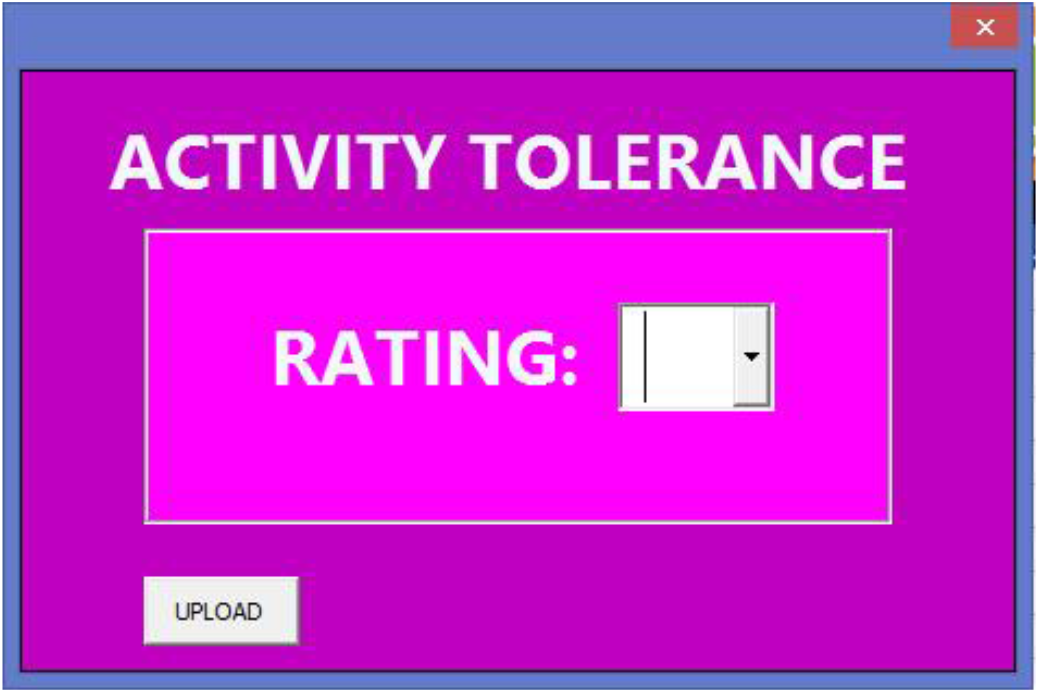
Activity Tolerance User Form

**Figure 3.3f.**
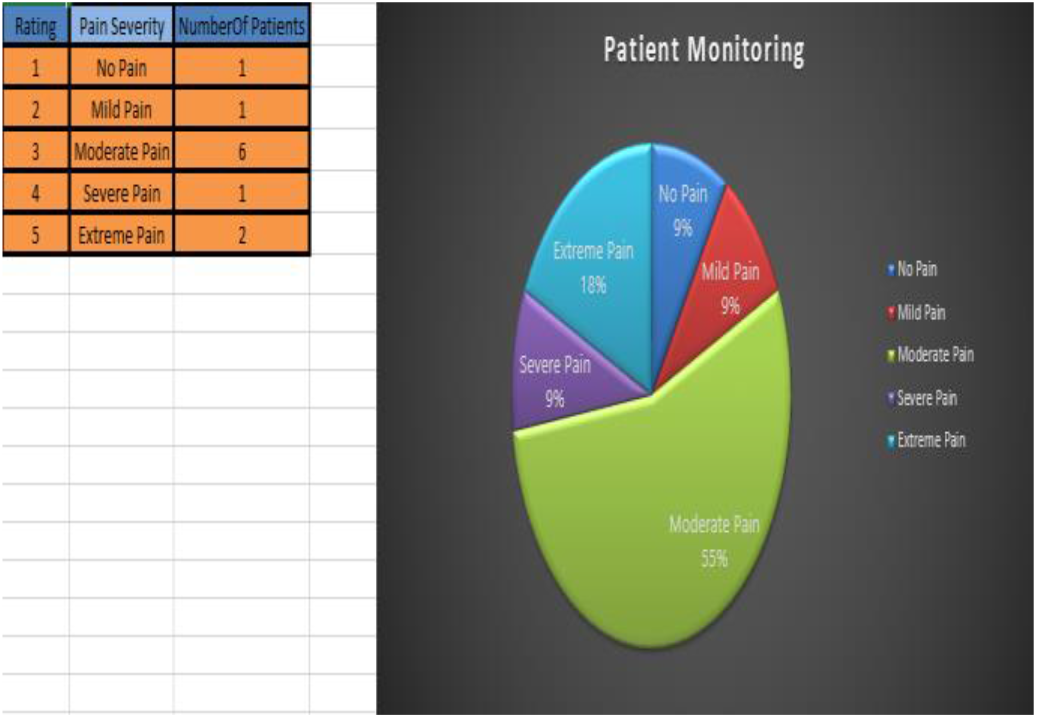
Patient Monitoring

**Figure 4.3a.**
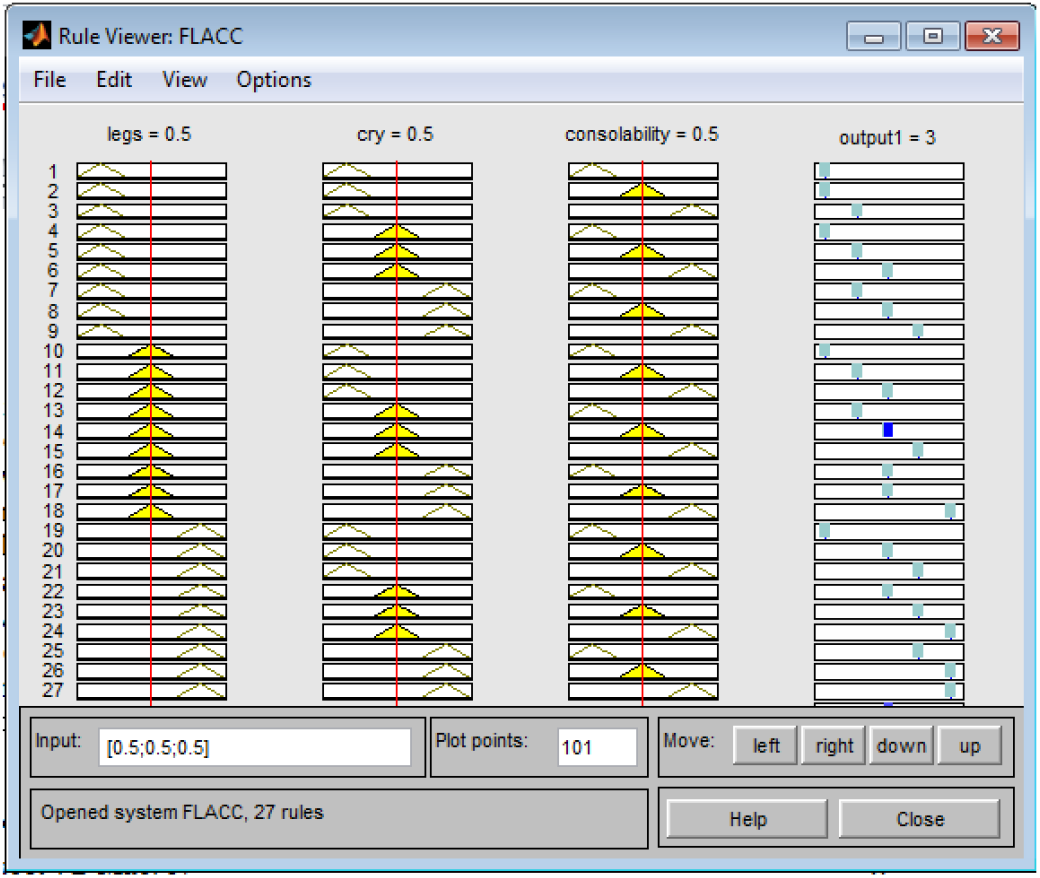
FLACC Simulation Results

**Figure 4.3b.**
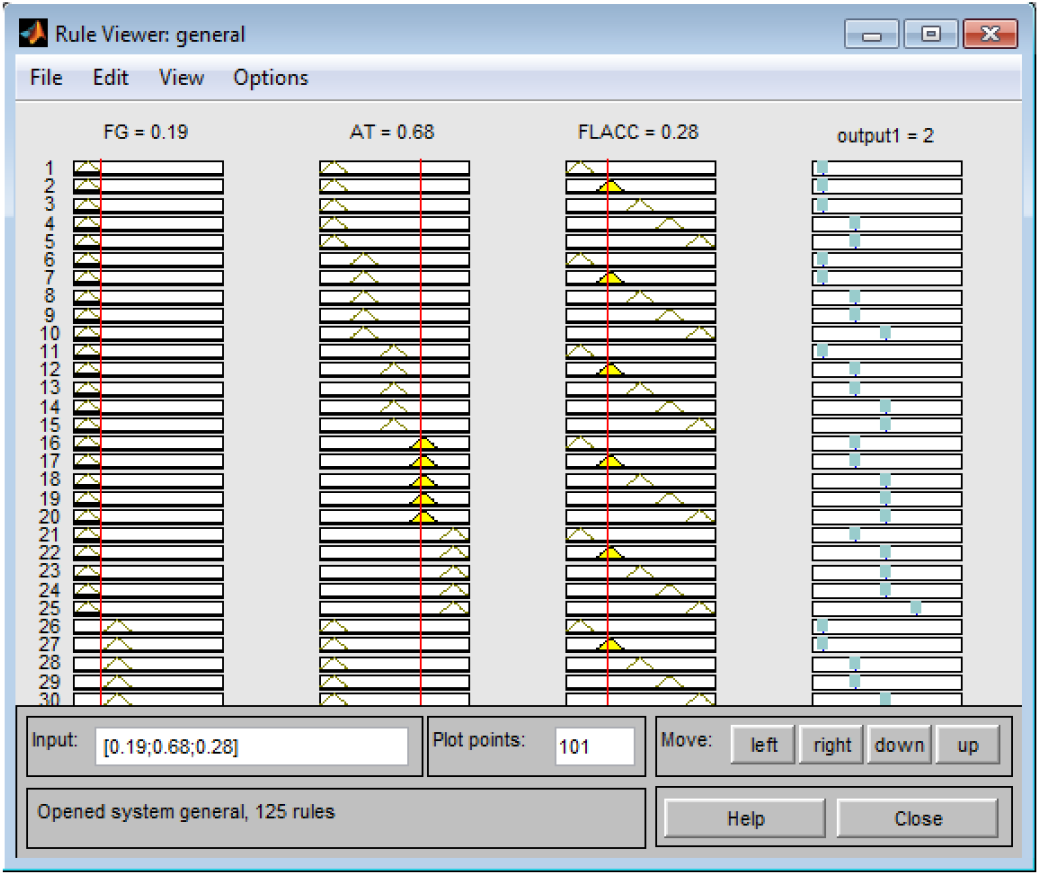
Pain Severity Simulation Results

After data has been encoded by the user, the program automatically generates the membership function and it calculates the input parameters. The sum of weights for each FAM matrix of pain parameter shall be summed up and the center of gravity shall be calculated. The crisp output shall be called *Pain Severity Assessment* and the linguistic term for obtained value shall remark. The update data values will be the reference of the chart for the purpose of monitoring. *See Figure 3.3f*

## 4. RESULTS AND DISCUSSIONS

### 1. Data Generation

The author proponents a number of different sets of experiments. Random data were inputted to yield and verify consistency in results. These were the compared to the results of MatLab. The data inputted are the values which are not normalized, the system was tasks to normalize it to adjust to its scale and program. The proponents have defined a marginal unit deduction to the input to avoid over lapping of values and to avoid error in the output.

### 2. Matlab Fuzzy Logic Toolbox

The Fuzzy Logic Toolbox™ is a powerful tool in a user-friendly environment that lets user model their complex system in a simplified structure through using simple logic rules and implementing these rules in a fuzzy inference system. The user is provided with different selections that would suit their system’s preference or requirement [9]. Below figures elicit the simulation results using Matlab Fuzzy Logic Toolbox.

### 4. Excel VB Macro Program

An Excel macro is an excel-based programming tool that entails set of instructions run by the user in their predefined keyboard shortcut, toolbar button or icon in a spreadsheet. It is a powerful tool used in different companies, which monitors the stability of the process on a trend chart. The code is written in Visual Basic for Applications (VBA) [9]. The proponents preferred to use this programming language as platform for the study since there is no tangible monitoring for each pain severity parameter. Excel macro, in this consideration, is equipped with statistical tools and charts that would address the need of visual monitoring and control of data.

### 5. Matlab Fuzzy Logic Toolbox versus Excel VB Macro Program

The results obtained using Matlab Fuzzy Logic Toolbox was compared with the results obtained using Excel VB Macro. The comparison between these two methods is shown in *tables 4.5a*. Having a perfect correlation, that is zero true error and relative approximate error, it was concluded that the results between the two methods have perfect correlation and therefore implies that the trials that was made have reliable and accurate results.

**Table 4.5a.** Pain Severity Parameters Assessment (Matlab Toolbox vs Excel VB Macro)

| TRIALS (based on data generated) | Pain Assessment Input Parameter | Input Values (Normalized) | Crisp Output (MATLAB Fuzzy Logic Toolbox) | Crisp Output (Excel VB Macro) | Linguistic Classification | True Error | % Relative Approximate Error |
| --- | --- | --- | --- | --- | --- | --- | --- |
| 1 | Facial Grimace | 0.1998 | 1 | 1 | NP | 0 | 0 |
|  | Activity Tolerance | 0.1998 |  |  |  |  |  |
|  | FLACC | 0.1998 |  |  |  |  |  |
| 2 | Facial Grimace | 0.7798 | 3 | 3 | MOP | 0 | 0 |
|  | Activity Tolerance | 0.5998 |  |  |  |  |  |
|  | FLACC | 0.5998 |  |  |  |  |  |
| 3 | Facial Grimace | 0.3998 | 2 | 2 | MP | 0 | 0 |
|  | Activity Tolerance | 0.3998 |  |  |  |  |  |
|  | FLACC | 0.5998 |  |  |  |  |  |
| 4 | Facial Grimace | 0.7998 | 5 | 5 | EP | 0 | 0 |
|  | Activity Tolerance | 0.7998 |  |  |  |  |  |
|  | FLACC | 0.9998 |  |  |  |  |  |
| 5 | Facial Grimace | 0.9998 | 5 | 5 | EP | 0 | 0 |
|  | Activity Tolerance | 0.5998 |  |  |  |  |  |
|  | FLACC | 0.9998 |  |  |  |  |  |
| 6 | Facial Grimace | 0.5998 | 3 | 3 | MOP | 0 | 0 |
|  | Activity Tolerance | 0.5998 |  |  |  |  |  |
|  | FLACC | 0.5998 |  |  |  |  |  |
| 7 | Facial Grimace | 0.5998 | 3 | 3 | MOP | 0 | 0 |
|  | Activity Tolerance | 0.7998 |  |  |  |  |  |
|  | FLACC | 0.5998 |  |  |  |  |  |
| 8 | Facial Grimace | 0.3998 | 3 | 3 | MOP | 0 | 0 |
|  | Activity Tolerance | 0.5998 |  |  |  |  |  |
|  | FLACC | 0.9998 |  |  |  |  |  |
| 9 | Facial Grimace | 0.7998 | 3 | 3 | MOP | 0 | 0 |
|  | Activity Tolerance | 0.5998 |  |  |  |  |  |
|  | FLACC | 0.5998 |  |  |  |  |  |
| 10 | Facial Grimace | 0.7998 | 3 | 3 | MOP | 0 | 0 |
|  | Activity Tolerance | 0.9998 |  |  |  |  |  |
|  | FLACC | 0.1998 |  |  |  |  |  |
| 11 | Facial Grimace | 0.9998 | 4 | 4 | SP | 0 | 0 |
|  | Activity Tolerance | 0.7998 |  |  |  |  |  |
|  | FLACC | 0.3998 |  |  |  |  |  |

## 5. CONCLUSIONS

The contribution of this paper is a tangible methodology for the decision-making involved in pain assessment for patients or users. The proposed system was implemented using Matlab Fuzzy Logic Toolbox and Excel VB Macro Program. The fuzzy system designed in these two platforms was successfully compiled and simulated. The results obtained from Excel VB Macro were compared with the results of MATLAB fuzzy logic toolbox, and exact correlation results were obtained. Most importantly, in this paper, the proponents provided a data-oriented fuzzy-based strategy applied to significant pain severity parameters derived from the existing pain assessment tools. The uncertainties involved in assessing the severity of pain was successfully addressed by comparing and analysing pain severity scale from different tools to come up with a generalized results.

## 6. RECOMMENDATIONS

In the absence of actual data, it is difficult to evaluate the pain severity in the context of medical and healthcare practice. The proposed system was designed for the purpose of assessing the pain severity and identifying its parameters and classification using fuzzy logic. The system was intended to integrate fuzzy logic in an Excel VB Macro program and test its reliability and accuracy through comparing it with MatLab Fuzzy Logic program. The output must also be compared to decisions made by experts to prove that the system can actually help in expert’s decision-making, otherwise the system will only be limited to self-report or self-assessment and therefore needs second opinion from the practitioner.

In addition, the proposed system was based from three specific pain assessment tools which are FLACC, Facial Grimace and Activity Tolerance Pain Scale, the proponents recommend the use of other existing pain assessment tools. It should also be noted that this study is unidimensional and only involves intensity of the pain from the patient’s point of view, it is recommended for the future researchers to conduct a similar study in pain assessment considering the multidimensional parameters of pain which includes social, sexual and cultural aspects of the patient.

## Data Availability

All data produced in the present study are available upon reasonable request to the authors

## ACKNOWLEDGMENTS

The authors acknowledges Jesus Christ, who is the source of wisdom and knowledge in the development of the fuzzy-based pain assessment system. They would like to thank Engr. Rionel Caldo, MPA for his expertise and professional advices as well as for encouraging us to pursue this study.

